# Factors associated with progression to critical illness in 28 days among COVID-19 patients: results from a tertiary care hospital in Istanbul, Turkey

**DOI:** 10.1101/2020.10.09.20209775

**Authors:** Uluhan Sili, Pinar Ay, Ahmet Topuzoglu, Huseyin Bilgin, Elif Tukenmez Tigen, Buket Erturk Sengel, Dilek Yagci Caglayik, Baran Balcan, Derya Kocakaya, Sehnaz Olgun Yildizeli, Fethi Gul, Beliz Bilgili, Rabia Can Sarinoglu, Aysegul Karahasan Yagci, Lutfiye Mulazimoglu Durmusoglu, Emel Eryuksel, Zekaver Odabasi, Haner Direskeneli, Sait Karakurt, Ismail Cinel, Volkan Korten

## Abstract

**Background:** Coronavirus disease 2019 (COVID-19) quickly spread worldwide to become a pandemic. This study aimed to define the predictors of critical illness development within 28 days postadmission.

**Methods:** We conducted a prospective cohort study including 477 PCR-positive COVID-19 patients admitted to a tertiary care hospital in Istanbul from March 12 to May 12, 2020. The development of critical illness, e.g., invasive mechanical ventilation and/or death, was followed for a period of 28 days postadmission. Demographic characteristics, number of comorbidities, illness severity at admission defined by the WHO scale, vital signs, laboratory findings and period of admission to the hospital were independent variables. Cox proportional hazards analysis was performed, and the C-index was calculated.

**Results:** The median (IQR) age of the cohort was 55.0 (44.0-67.0) years, and 50.1% were male. The most common presenting symptoms were cough, dyspnea and fatigue. Overall, 65.2% of the patients had at least one comorbidity. Hydroxychloroquine was given to 99.2% of the patients. Critical illness developed in 45 (9.4%; 95% CI: 7.0%-12.4%) patients. In the multivariable analysis, age (HR: 1.05, p<0.001), number of comorbidities (HR: 1.33, p=0.02), procalcitonin ≥0.25 µg/L (HR: 2.12, p=0.03) and LDH ≥350 U/L (HR: 2.04, p=0.03) were independently associated with critical illness development. The WHO scale on admission was the strongest predictor of critical illness (HR: 4.15, p<0.001). Prognosis improved within the study period (p<0.05). The C-index of the model was 0.92.

**Conclusions:** Age, comorbidity number, WHO scale, LDH and procalcitonin were independently associated with critical illness development. Mortality from COVID-19 seems to be decreasing as the pandemic advances.

**summary:** We analyzed 477 confirmed COVID-19 patients for the development of critical illness, e.g., invasive mechanical ventilation and/or death within 28 days postadmission. Age, comorbidity number, WHO scale, LDH and procalcitonin were independently associated with critical illness development.

## INTRODUCTION

The first coronavirus disease 2019 (COVID-19) case in Turkey was diagnosed on 11 March 2020.^1^ As of 17 August 2020, a total of 249309 cases were confirmed with 5974 deaths. Although the daily case numbers have declined, approximately 1000 new cases are still diagnosed every day. Istanbul, a metropolis of approximately 16 million inhabitants, is the epicenter of COVID-19 in Turkey.

The pandemic response in Turkey was led by a scientific council formed by the government. Hospitals acted according to the regularly updated guidelines published by the scientific council for the diagnosis and treatment of COVID-19 patients. Admission to the hospital was recommended for patients over the age of 50 years and those with comorbid conditions, severe pneumonia, and/or laboratory values indicative of worse prognosis. All patients hospitalized with COVID-19 pneumonia were given empirical hydroxychloroquine treatment per the council’s recommendation.^2^ There is limited information on the prognosis of COVID-19 patients in Turkey.

COVID-19 can progress differently among infected individuals, ranging from asymptomatic carriage to respiratory failure leading to death.^3^ In a report of 72314 cases from China, the spectrum of disease varied from mild (81%) and severe (14%) to critical (5%).^4^ In a recent systemic review and meta-analysis, the all-cause mortality rate of hospitalized COVID-19 patients was 10%, with a predictive interval of 2% to 39%.^5^ The authors concluded that there was substantial between-study heterogeneity, which led to a large 95% predictive interval suggesting high uncertainty. Clearly, high-quality cohort studies with defined severity criteria and adequate follow-up times are needed.^6, 7^ It is important to determine the critical illness outcomes and associated factors, which are vital to determine hospitalization criteria, which in turn will affect medical resource allocation.^8^

Here, we describe the demographic characteristics, clinical features, 28-day critical illness development and factors associated with critical illness among COVID-19 patients treated in one of the main pandemic hospitals in Istanbul.

## METHOD

### Study setting and design

We conducted a single-center, prospective cohort study. All hospitalized ≥18-year-old patients with confirmed COVID-19, i.e., positive RT-PCR assay for the SARS-CoV-2 genome detected in a nasopharyngeal and/or oropharyngeal swab specimen were included. The study period started with the first confirmed patient in our hospital on March 12, 2020, and included patients hospitalized until May 12, 2020. Testing was performed on patients who fulfilled the criteria for possible COVID-19 according to the Turkish COVID-19 guideline.^2^ Essentially, patients whose symptoms and signs were compatible with viral pneumonia (e.g., fever, cough, dyspnea or fatigue) and some cases based on clinical judgment were tested. The decision to admit a suspected COVID-19 patient to the hospital was mostly a clinical decision based on age, presence of comorbidity, dyspnea, hypoxia and extent of pulmonary involvement detected with thoracic CT.

### Study participants

There were 521 patients hospitalized within the study period. The patients (n=55) who were transferred to or from our center were included in the study provided that their medical records could be retrieved from other centers. As the outcome was the development of critical illness, we excluded patients who had invasive mechanical ventilation and/or death within 24 hours of admission (n=41) (Figure 1). Healthcare-associated COVID-19 cases were also excluded (n=3). After these exclusions, 477 patients were eligible and followed-up for a period of 28 days postadmission.

**Figure 1.**
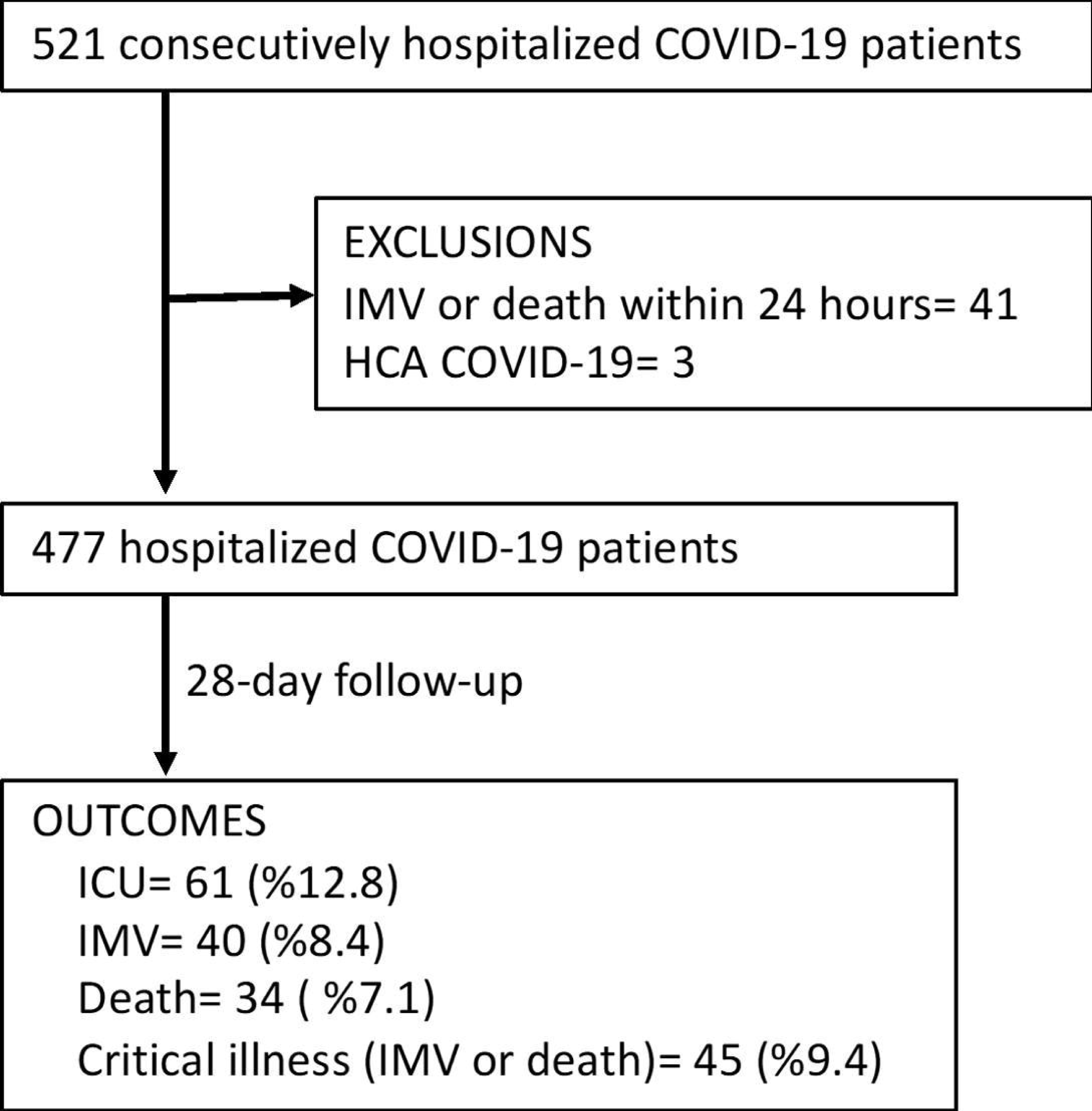
Flowchart of included patients and their outcomes. ICU, intensive care unit; IMV, invasive mechanical ventilation; HCA, healthcare associated.

### Data collection

Demographic information, exposure history, comorbidities, comedications, symptoms, signs, imaging and PCR results, vital signs and clinical course including medications, response to treatments, laboratory parameters, intensive care unit (ICU) referral and prognosis were prospectively collected in forms standardized for COVID-19 patients. This information was then transferred to an electronic database, after which four physicians (US, PA, AT, HB) went through the data extensively, cross-checked the authenticity and completed missing information using archival and electronic records.

### Definition of the variables

The outcome was progression to critical illness, which was either invasive mechanical ventilation or death within 28 days postadmission. To determine readmission status, the patients were reached by a phone call, and survival status was checked from a centralized electronic death registration system.

The illness severity of the patients on the day of hospitalization was defined by the World Health Organization (WHO) ordinal scale for clinical improvement.^9^ This is an 8-point ordinal scale including 0, uninfected; 1, ambulatory with no limitation of activities; 2, ambulatory with limitation of activities; 3, hospitalized, mild disease, no oxygen therapy; 4, hospitalized, mild disease, oxygen by mask or nasal prongs; 5, hospitalized, severe disease, noninvasive ventilation or high-flow oxygen; 6, hospitalized, severe disease, intubation and mechanical ventilation; 7, hospitalized, severe disease, ventilation and additional organ support such as renal replacement therapy or extracorporeal membrane oxygenation; and 8, death.

Although the minimum scale for hospitalization was 3, we had some patients with a score of 2 who could have been followed in an ambulatory setting.

Age, sex, smoking status, body mass index (BMI), number of comorbidities, vital signs, laboratory findings and period of admission to the hospital were the independent variables. For vital signs, the worst recorded value within the first 24 hours of hospitalization was recorded. The quick sepsis-related organ dysfunction assessment (qSOFA) score was also calculated.^10^ The first available laboratory findings within 72 hours of hospital admission were noted. The period of admission to the hospital was categorized as two weekly periods within the study time.

### Statistics

Descriptive statistics are presented as percentages and medians (IQRs). Categorical variables were compared with the chi-square and Fisher’s exact tests. Continuous variables for two independent groups were analyzed by the Mann-Whitney U test. Multivariable analysis was carried out with 451 (94.5%) of the 477 patients due to missing data. Cox proportional hazards with stepwise backward elimination was used, and the strengths of associations are presented as hazard ratios (HRs). Incidence rates and HRs were presented with 95% confidence intervals (CIs). The concordance index (C-index) was calculated. P<0.05 was considered statistically significant.

### Approval

The study was approved by the ethics board of Marmara University, School of Medicine (09.2020.572). The requirement for written informed consent was waived by the board. We followed the Strengthening Reporting of Observational Studies in Epidemiology (STROBE) guidelines.

## RESULTS

Overall, 477 patients hospitalized with confirmed COVID-19 were included in the analysis. The median (IQR) age of the cohort was 55.0 (44.0-67.0) years, and 239 (50.1%) were male. An exposure history with a possible/definitive COVID-19 case was present in 229 (48.0%) patients.

The most common presenting symptom was cough followed by dyspnea, fatigue, myalgia, fever and headache (Table 1). All symptoms except dyspnea, diarrhea, rhinorrhea and confusion were significantly more common in patients younger than 60 years than in elderly patients (p<0.05 for all).

**Table 1.** Frequency of symptoms by age.

|  | All patients |  | <60 years |  | ≥60 years |  | p value |
| --- | --- | --- | --- | --- | --- | --- | --- |
|  | n | % | n | % | n | % |  |
| Cough | 348 | 73.0 | 216 | 76.6 | 132 | 67.7 | 0.031 |
| Dyspnea | 270 | 56.6 | 155 | 55.0 | 115 | 59.0 | >0.05 |
| Fatigue | 270 | 56.6 | 174 | 61.7 | 96 | 49.2 | 0.007 |
| Myalgia | 202 | 42.3 | 138 | 48.9 | 64 | 32.8 | <0.001 |
| Fever | 201 | 42.1 | 133 | 47.2 | 68 | 34.9 | 0.008 |
| Headache | 165 | 34.6 | 113 | 40.1 | 52 | 26.7 | 0.004 |
| Nausea | 120 | 25.2 | 87 | 30.9 | 33 | 16.9 | 0.001 |
| Anosmia/ageusia | 93 | 19.5 | 72 | 25.6 | 21 | 10.8 | <0.001 |
| Sore throat | 87 | 18.2 | 61 | 21.6 | 26 | 13.3 | 0.021 |
| Diarrhea | 68 | 14.3 | 43 | 15.2 | 25 | 12.8 | >0.05 |
| Vomiting | 57 | 12.0 | 43 | 15.3 | 14 | 7.2 | 0.007 |
| Rhinorrhea | 33 | 6.9 | 23 | 8.2 | 10 | 5.1 | >0.05 |
| Confusion | 7 | 1.5 | 4 | 1.4 | 3 | 1.5 | >0.05 |

Sociodemographic variables, comorbidities, smoking status, BMI, vital signs, laboratory findings and the period of hospital admission are presented in Table 2. At admission, 354 (74.2%) cases were categorized as WHO scale 2&3, while 123 (25.8%) were categorized as WHO scale 4&5.

**Table 2.** Clinical characteristics on admission associated with progression to critical illness within 28 days of follow-up.

|  | All patients |  | Did not progress to critical illness |  | Progressed to critical illness |  |  |
| --- | --- | --- | --- | --- | --- | --- | --- |
| Variable | (n=477) |  | (n=432) |  | (n=45) |  | p value |
|  | n | % | n | % | n | % |  |
| Age (years) |  |  |  |  |  |  |  |
| ≤39 | 87 | 18.2 | 87 | 20.1 | 0 | 0.0 |  |
| 40 - 59 | 195 | 40.9 | 185 | 42.8 | 10 | 22.2 |  |
| 60 - 79 | 153 | 32.1 | 127 | 29.4 | 26 | 57.8 |  |
| ≥80 | 42 | 8.8 | 33 | 7.6 | 9 | 20.0 | <0.001 |
| Sex |  |  |  |  |  |  |  |
| Male | 239 | 50.1 | 212 | 49.1 | 27 | 60.0 | >0.05 |
| Comorbidity |  |  |  |  |  |  |  |
| Hypertension | 220 | 46.1 | 188 | 43.5 | 32 | 71.1 | <0.001 |
| Diabetes | 139 | 29.1 | 120 | 27.8 | 19 | 42.2 | 0.042 |
| Cardiovascular disease<sup>1</sup> | 109 | 22.9 | 88 | 20.4 | 21 | 46.7 | <0.001 |
| Chronic lung disease<sup>2</sup> | 87 | 18.2 | 71 | 16.4 | 16 | 35.6 | 0.002 |
| Neuropsychiatric condition<sup>3</sup> | 41 | 8.6 | 31 | 7.2 | 10 | 22.2 | 0.003 |
| Immune compromised state<sup>4</sup> | 34 | 7.1 | 26 | 6.0 | 8 | 17.8 | 0.009 |
| Chronic kidney disease | 29 | 6.1 | 24 | 5.6 | 5 | 11.1 | >0.05 |
| Chronic liver disease | 6 | 1.3 | 5 | 1.2 | 1 | 2.2 | >0.05 |
| ≥1 comorbidity | 311 | 65.2 | 271 | 62.7 | 40 | 88.9 | <0.001 |
| Smoking status |  |  |  |  |  |  |  |
| Current | 20 | 4.2 | 17 | 3.9 | 3 | 6.7 |  |
| Former | 78 | 16.4 | 70 | 16.2 | 8 | 17.8 |  |
| Never | 379 | 79.5 | 345 | 79.9 | 34 | 75.6 | >0.05 |
| Body mass index<sup>5</sup> |  |  |  |  |  |  |  |
| ≤24.9 | 95 | 23.2 | 85 | 22.9 | 10 | 25.6 |  |
| 25.0 - 29.9 | 159 | 38.8 | 143 | 38.5 | 14 | 41.0 |  |
| ≥30.0 | 156 | 38.0 | 143 | 38.5 | 13 | 33.3 | >0.05 |
| Time from illness onset to hospitalization, median (IQR) | 5 (3.0-8.0) |  | 5.0 (3.0-8.0) |  | 6.5 (2.0-9.5) |  | >0.05 |
| Vital signs |  |  |  |  |  |  |  |
| Temperature ≥38°C | 91 | 19.2 | 76 | 17.6 | 15 | 34.9 | 0.006 |
| Heart rate >90 beats/min | 268 | 56.5 | 238 | 55.2 | 30 | 69.8 | 0.067 |
| Systolic BP ≤100 mmHg | 74 | 15.6 | 67 | 15.5 | 7 | 16.7 | >0.05 |
| Mean arterial BP <65 mmHg | 8 | 1.7 | 6 | 1.4 | 2 | 4.8 | >0.05 |
| Respiratory rate ≥22/min | 304 | 64.1 | 268 | 62.5 | 36 | 80.0 | 0.020 |
| Oxygen saturation ≤93% | 157 | 33.1 | 124 | 28.8 | 33 | 75.0 | <0.001 |
| qSOFA >1 | 61 | 12.9 | 50 | 11.6 | 11 | 26.2 | 0.007 |
| WHO scale on admission |  |  |  |  |  |  |  |
| WHO-2 | 58 | 12.2 | 58 | 13.4 | 0 | 0.0 |  |
| WHO-3 | 296 | 62.1 | 287 | 66.4 | 9 | 20.0 |  |
| WHO-4 | 119 | 24.9 | 86 | 19.9 | 33 | 73.3 |  |
| WHO-5 | 4 | 0.8 | 1 | 0.2 | 3 | 6.7 | <0.001 |
| Laboratory findings |  |  |  |  |  |  |  |
| Lymphocyte <800/ mm<sup>3</sup> | 89 | 18.9 | 67 | 15.7 | 22 | 48.9 | <0.001 |
| CRP ≥30 mg/dL | 218 | 46.8 | 177 | 42.0 | 41 | 91.1 | <0.001 |
| Procalcitonin ≥0.25 µg/L | 21 | 11.0 | 30 | 7.1 | 21 | 46.7 | <0.001 |
| Ferritin >300 µg/L | 126 | 27.8 | 97 | 23.8 | 29 | 64,4 | <0.001 |
| D-dimer ≥1.0 mg/L | 128 | 27.4 | 99 | 23.3 | 29 | 67.4 | <0.001 |
| Fibrinogen >400 mg/dL | 264 | 64.2 | 233 | 62.0 | 31 | 88.6 | 0.002 |
| LDH $\geq 350$ U/L | 108 | 23.0 | 84 | 19.8 | 24 | 53.3 | <0.001 |
| Troponin T >14 ng/L | 89 | 22.4 | 66 | 18.2 | 23 | 65.7 | <0.001 |
| Period of admission to hospital |  |  |  |  |  |  |  |
| 12-27 March | 31 | 6.5 | 22 | 5.1 | 9 | 20.0 |  |
| 28 March-11 April | 208 | 43.6 | 190 | 44.0 | 18 | 40.0 |  |
| 12-26 April | 158 | 33.1 | 142 | 32.9 | 16 | 35.6 |  |
| 27 April-12 May | 80 | 18.8 | 78 | 18.1 | 2 | 4.4 | <0.001 |

Abbreviations: BP, blood pressure; CRP, C-reactive protein; LDH, lactate dehydrogenase; qSOFA, quick sepsis-related organ dysfunction assessment;
<sup>1</sup>excluding hypertension.
<sup>2</sup>includes asthma, chronic obstructive pulmonary disease, bronchiectasis and interstitial pulmonary fibrosis.
<sup>3</sup>includes dementia, cerebrovascular disease and psychiatric conditions.
<sup>4</sup>includes rheumatologic disorders, immunosuppressive treatment, recent diagnosis of hematologic/solid organ malignancy.
<sup>5</sup>BMI was missing in 67 (14.0%) participants.

Among the 477 patients, 61 (12.8%, 95%CI: 10.0%-16.1%) were admitted to the ICU. Forty patients (8.4%, 95%CI: 6.1%-11.2%) were intubated and mechanically ventilated. Thirty-four (7.1%, 95%CI: 5.0%-9.8%) patients died within the 28-day follow-up (Figure 1). Overall, 45 (9.4%, 95% CI: 7.0%-12.4%) patients developed critical illness. The median (IQR) duration from hospital admission to invasive mechanical ventilation and death was 5.0 (3.0 – 8.8) and 12.0 (8.0 – 20.0) days, respectively. The median (IQR) hospital length of stay was 7.0 (5.0 – 11.0) days.

Hydroxychloroquine alone and combined with azithromycin were administered to 473 (99.2%) and 199 (41.7%) patients, respectively. Favipiravir (n=147, 30.8%) was used in patients with peripheral oxygen saturation ≤93% in ambient air. Oseltamivir was coadministered to 31 (6.5%) patients. Corticosteroid and tocilizumab were given to 46 (9.6%) and 30 (6.3%) patients, respectively. Anticoagulant thromboprophylaxis with low-molecular-weight heparin (LMWH) was given to 295 (61.8%) patients. While 14 (45.2%) patients were given LMWH prophylaxis in the first two weeks of the study period, this number increased to 62 (77.5%) in the last two weeks (p<0.001).

Patients who progressed to critical illness were older and more likely to have comorbid conditions such as hypertension, diabetes, cardiovascular disease and chronic lung disease (Table 2). The development of critical illness was not associated with smoking status or BMI (p>0.05 for both). Patients who had progressed to critical illness were more likely to have a temperature ≥38°C, tachypnea and hypoxia at the time of hospital admission. Significantly more patients with qSOFA >1 progressed to critical illness than those with qSOFA ≤1. Patients progressing to critical illness had significantly higher rates of lymphopenia and higher C-reactive protein (CRP), procalcitonin, ferritin, d-dimer, fibrinogen, lactate dehydrogenase (LDH), and troponin T levels (Table 2).

In patients younger than 60 years old, the presence of comorbidity was a significant factor for the development of critical illness. Only one (0.7%) patient younger than 60 without comorbidity developed critical illness compared to 9 (6.2%) with comorbidity (p=0.02).

The WHO scale for clinical improvement showed a significant association with the progression to critical illness. While 29.3% of patients with WHO scale 4&5 on admission progressed to critical illness, this rate was 2.5% for those with WHO scale 2&3 (p<0.001) (Figure 2). The rate of critical illness decreased significantly as the pandemic advanced. While 29.0% of patients who were hospitalized during the first two weeks of the pandemic progressed to critical illness, this rate was 2.5% for patients hospitalized during the last two weeks of the study period (p<0.001) (Figure 3).

**Figure 2.**
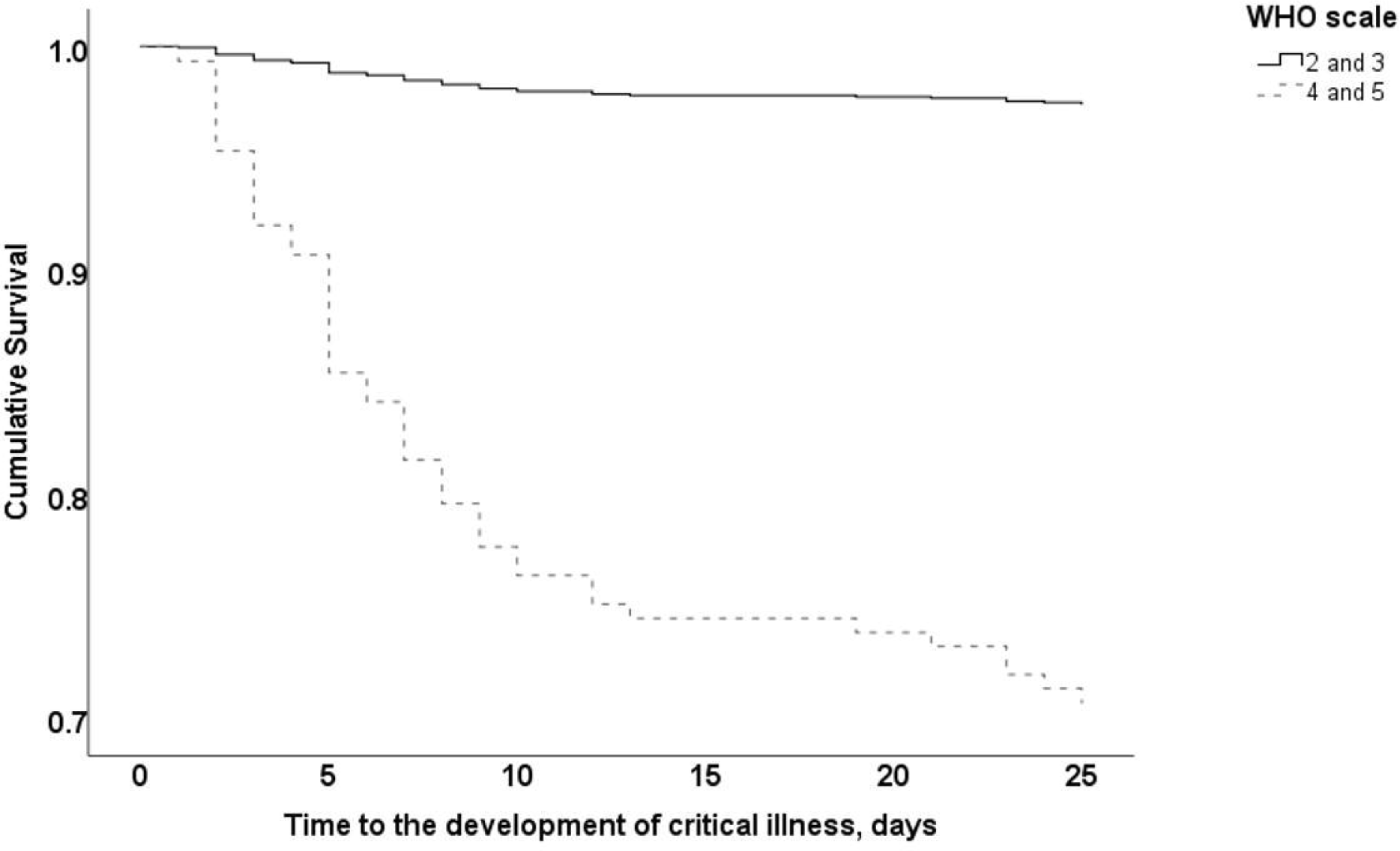
Time to development of critical illness by the baseline WHO scale for clinical improvement.

**Figure 3.**
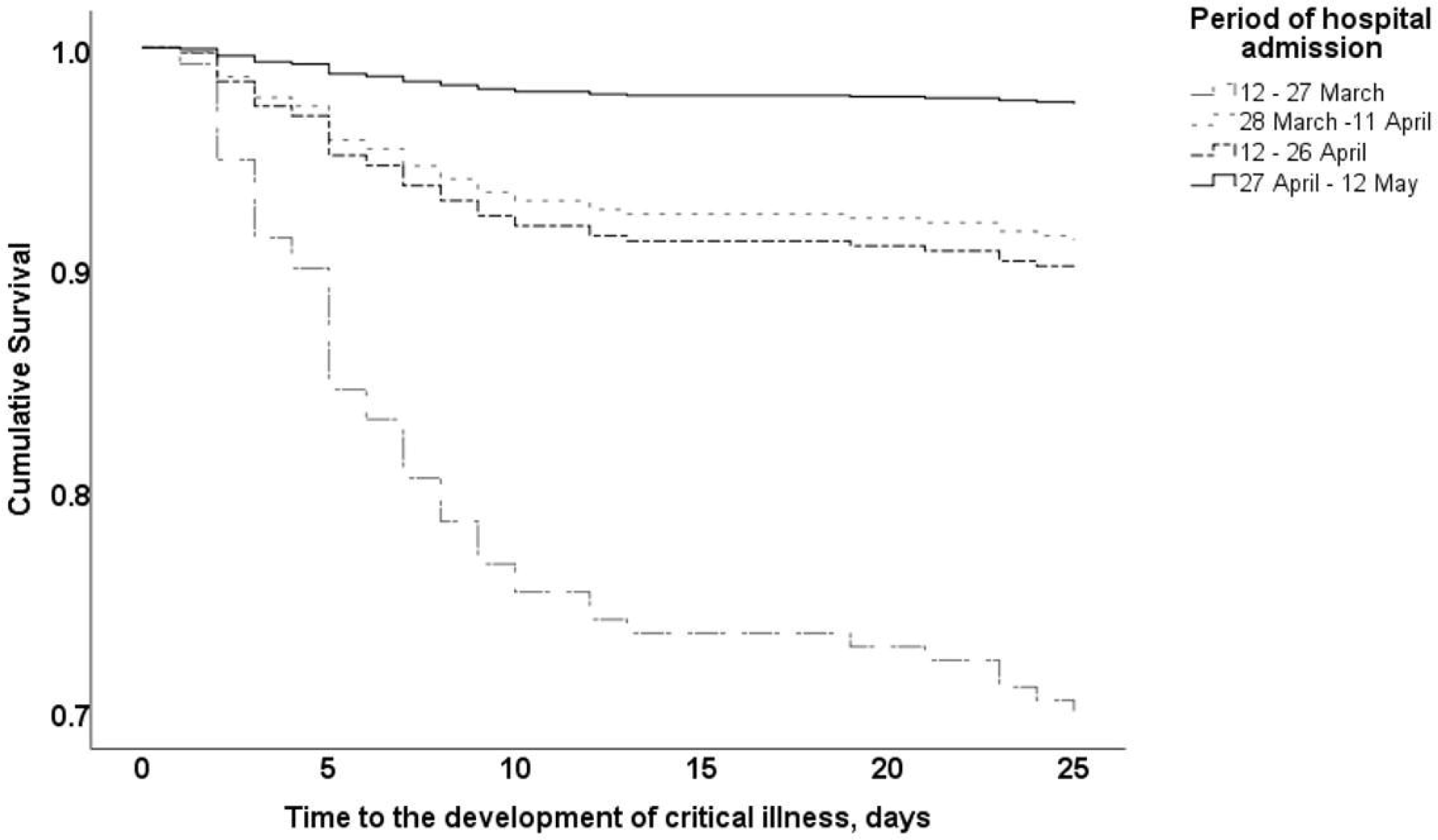
Time to development of critical illness by the period of hospital admission.

Univariable and multivariable analyses for progression to critical illness within the 28-day follow-up are presented in Table 3. Age, sex, number of comorbidities, WHO scale for clinical improvement, baseline laboratory values and period of admission to hospital were included in the multivariable model. Age was significantly associated with progression to critical illness; each year had an HR of 1.05 (95%CI: 1.02-1.08). The number of comorbidities was also independently associated with critical illness (HR: 1.33, 95% CI: 1.05-1.68). Among the laboratory values, procalcitonin ≥0.25 µg/L (HR: 2.12, 95%CI: 1.07-4.21) and LDH ≥350 U/L (HR: 2.04, 95%CI: 1.06-3.94) were independently associated with critical illness. At admission, WHO scale of 4&5 had an HR of 4.15 (95%CI: 1.87-9.22) for progression to critical illness compared to WHO scale of 2&3. The risk of progression to critical illness decreased significantly as the hospitalization period advanced. Compared to the first period, the HR was 0.14 (95%CI: 0.03-0.72) in patients hospitalized during the last two weeks. The C-index of the model was 0.92.

**Table 3.** Cox regression analysis for association between clinical characteristics and progression to critical illness within 28 days of follow-up.

|  | Univariable analysis |  | Multivariable analysis |  |
| --- | --- | --- | --- | --- |
|  | HR (95%CI) | p value | HR (95%CI) | p value |
| Age | 1.06 (1.04-1.08) | <0.001 | 1.05 (1.02-1.08) | <0.001 |
| Period of hospital admission |  |  |  |  |
| 12-27 March | 1.00 |  | 1.00 |  |
| 28 March-11 April | 0.25 (0.11-0.57) | 0.001 | 0.38 (0.15-0.94) | 0.04 |
| 12-26 April | 0.30 (0.13-0.67) | 0.004 | 0.68 (0.27-1.75) | 0.43 |
| 27 April-12 May | 0.07 (0.02-0.33) | 0.001 | 0.14 (0.03-0.72) | 0.02 |
| Lymphocyte <800/ mm<sup>3</sup> | 4.53 (2.52-8.12) | <0.001 | 1.93 (1.00-3.75) | 0.05 |
| CRP ≥30 mg/dL | 12.80 (4.58-35.73) | <0.001 | 3.07 (1.00-9.44) | 0.05 |
| Procalcitonin ≥0.25 µg/L | 8.82 (4.90-15.86) | <0.001 | 2.12 (1.07-4.21) | 0.03 |
| D-dimer ≥1.0 mg/L | 6.02 (3.18-11.39) | <0.001 | - |  |
| LDH ≥350 U/L | 4.26 (2.37-7.65) | <0.001 | 2.04 (1.06-3.94) | 0.03 |
| Number of co-morbidities | 1.79 (1.46-2.20) | <0.001 | 1.33 (1.05-1.68) | 0.02 |
| WHO scale on admission |  |  |  |  |
| 2&3 | 1.00 |  | 1.00 |  |
| 4&5 | 13.32 (6.41-27.66) | <0.001 | 4.15 (1.87-9.22) | <0.001 |

Abbreviations: CRP, C-reactive protein; LDH, lactate dehydrogenase.

## DISCUSSION

This study describes the demographic characteristics, clinical features and prognosis of 477 COVID-19 patients followed by a university hospital in Istanbul from mid-March to mid-May. The median age was 55.0 years (IQR 44.0-67.0), with an equal distribution of sexes. The 28-day mortality rate was 7.1%. The rate of progression to critical illness was 9.4%.

Some studies with comparable characteristics of participants reported similar rates for the development of critical illness or death. A cohort study from China including patients with a mean age of 46.1 years reported a rate of 8% for the development of adverse outcomes.^11^ Another study from two hospitals in New York City including patients with a median age of 62.2 years revealed a mortality rate of 10.2%.^12^ Comparing these rates could be misleading due to the differences in the testing criteria influencing case identification, hospitalization indications and case mix across studies. Additionally, while some of the cohorts evaluated in-hospitality mortality, others focused on 14- or 28-day survival rates, which make comparisons even more problematic. Nevertheless, our rates for the development of critical illness seem somewhat lower than those of most of the studies, as Wiersinga et al. indicate that the overall hospital mortality from COVID-19 is approximately 15%–20%.^3^ A major difference in our cohort was the extensive use of hydroxychloroquine. Whether hydroxychloroquine administration lowered the rates of critical illness development is beyond the scope of this study.^13^

The presenting symptoms were similar to those reported in other studies; cough, dyspnea and fatigue were the most common^3^, and one-fifth of the patients reported anosmia and/or ageusia.^14, 15^ Almost all symptoms were more common among patients <60 years than among elderly patients. This might be due to a predilection to hospitalize younger patients with more pronounced symptoms. This finding also suggests that presenting complaints might be subtler among elderly patients, as clinical presentation might vary with age.^15^ Sixty-five percent of the patients had at least one comorbidity, such as hypertension, diabetes or cardiovascular disease, which is consistent with other studies’ findings.^3^

Univariable analysis revealed some risk factors for critical illness that were already described in the literature.^3, 6, 16-18^ Older age, comorbidities, fever, tachypnea and hypoxia at presentation were associated with progression to critical illness. Laboratory abnormalities such as lymphopenia, elevated acute phase reactants and coagulation abnormalities were also associated with critical illness. Contrary to the findings of Zheng et al., smoking status was not associated with critical illness development.^19^ This is probably due to a reporting bias, as the 4.2% current smoker rate in our cohort is much lower than the 28.0% daily smoker rate in Turkey.^20^ While Petrilli et al. found BMI >40 to be independently associated with critical illness development, our findings did not reveal such an association.^17^ In our cohort, BMI was calculated based on self-report of the patients and was missing in 67 (14%) participants.

Multivariable analysis showed that age was independently associated with critical illness, as reported by other studies.^3, 21^ The number of comorbidities was also associated with critical illness; each comorbidity increased the HR by 1.33. Liang et al. also found the number of comorbidities as a significant predictor for the development of critical illness.^22^ We did not enter specific comorbidities into the model testing, as comorbidities blend and cancel each other out during multivariable analysis. Our model has a high discrimination power with a C-index of 0.92.

The Turkish guidelines for the management of COVID-19 recommend hospitalization for COVID-19 patients >50 years regardless of comorbidity.^2^ Our analysis showed that for patients <60 years, comorbidity is required for progression to critical illness. This result suggests that the lower age limit of hospitalization for patients without comorbidity could be raised to 60. Such a change would relieve the demand for hospital beds.

Patients with PCT ≥0.25 µg/L and LDH ≥350 U/L had HRs of 2.12 and 2.04, respectively, for critical illness. PCT increases are expected in bacterial infections, suggesting that these patients might have coexisting bacterial infections.^17^ The LDH increase is probably due to cellular damage induced by the virus as well as the immune response against it. PCT and LDH associations were also reported in other studies.^17, 23, 24^ Although lymphopenia (<800/mm^3^) and CRP (≥30 mg/dL) remained in the model, they had borderline significance (p=0.05 for both). D-dimer (≥1 mg/L) did not remain in the model. As d-dimer levels correlated with CRP and PCT, it was omitted from the model at the last step of multivariable analysis.

The WHO scale for clinical improvement was significantly associated with progression to critical illness. Patients with a WHO scale of 4&5 had an HR of 4.15 for critical illness compared to those with WHO 2&3. The major difference between the WHO 4&5 and 2&3 scales is the need for supplemental oxygen. We chose to use the WHO scale in our multivariable model rather than qSOFA because it considers the need for oxygen supplementation directly. Additionally, the WHO scale showed a stronger association with critical illness than the qSOFA score. To the best of our knowledge, one other study from France also found the WHO scale category to be independently associated with the development of critical illness.^25^ Due to the limited number of patients, we could not test WHO scale categories separately in the multivariable model.

One factor that strongly remained in the model was the period of admission to the hospital. Patients admitted to our hospital at the beginning of May had an HR of 0.14 for progression to critical illness compared to patients admitted at the beginning of the pandemic in mid-March. Such an association was also reported in New York; patients admitted at the beginning of April had an OR of 0.08 for the risk of critical illness compared to patients admitted at the beginning of March.^17^ There could be several explanations for this observation. Our hospitalization criteria did not change as the pandemic progressed. Hospital care might have become more efficient as experience was gained with time. Medication protocols were slightly altered. Venous thromboprophylaxis using LMWH was initially administered to patients with classic risk factors for venous thrombosis; however, as coagulopathy became apparent in COVID-19 pathogenesis, more patients hospitalized with suspected or proven COVID-19 received prophylactic LMWH at the beginning of hospitalization unless contraindicated.^26^ These hypotheses warrant further investigation. Additionally, a less virulent form of the virus might have emerged, as there are reports showing variants of SARS-CoV-2 with differing infectivity and virulence from March to April.^27, 28^ Furthermore, patients might have been less exposed to the virus due to the increased utilization of preventive and control measures as the pandemic advanced.

Our study has several limitations. Although our hospital is one of the main pandemic hospitals within the region, this study is based on a single center’s experience. BMI information was lacking in 14.0% of the participants. We did not quantify the extent of pulmonary involvement on thoracic CT, which might predict the development of critical illness.

## CONCLUSIONS

Age, comorbidity number, WHO scale, LDH (≥350 U/L) and procalcitonin (≥0.25 µg/L) are independently associated with critical illness development in hospitalized COVID-19 patients. Mortality from COVID-19 seems to be decreasing as the pandemic advances. Our findings will be useful in shaping the response for the expected waves of COVID-19.

## Data Availability

The data that support the findings of this study are available on request from the corresponding authors [US and VK]. The data are not publicly available due to them containing information that could compromise research participant privacy.

## ACKNOWLEDGEMENTS

None of the authors report any conflict of interest relevant to this study. This manuscript was finalized on August 17, 2020.

## REFERENCES

1. Republic of Turkey Ministry of Health. COVID-19 Weekly Situation Report 13/07/2020 – 19/07/2020, https://dosyamerkez.saglik.gov.tr/Eklenti/38021,covid-19-weekly-situation-report---29-weekpdf.pdf?0&_tag1=44DBE7E25D380A645A4780C04D8C19C0EF692249 (accessed 17 August 2020).

2. Republic of Turkey Ministry of Health. COVID-19 Rehberi (Guideline), https://covid19bilgi.saglik.gov.tr/tr/covid-19-rehberi.html (accessed 17 August 2020).

3. Wiersinga WJ, Rhodes A, Cheng AC, et al. Pathophysiology, Transmission, Diagnosis, and Treatment of Coronavirus Disease 2019 (COVID-19): A Review. JAMA 2020 2020/07/11. DOI: 10.1001/jama.2020.12839.

4. Wu Z and McGoogan JM. Characteristics of and Important Lessons From the Coronavirus Disease 2019 (COVID-19) Outbreak in China: Summary of a Report of 72314 Cases From the Chinese Center for Disease Control and Prevention. JAMA 2020 2020/02/25. DOI: 10.1001/jama.2020.2648.

5. Potere N, Valeriani E, Candeloro M, et al. Acute complications and mortality in hospitalized patients with coronavirus disease 2019: a systematic review and meta-analysis. Crit Care 2020; 24: 389. 2020/07/04. DOI: 10.1186/s13054-020-03022-1.

6. Wynants L, Van Calster B, Collins GS, et al. Prediction models for diagnosis and prognosis of covid-19 infection: systematic review and critical appraisal. BMJ 2020; 369: m1328. 2020/04/09. DOI: 10.1136/bmj.m1328.

7. Anesi GL, Halpern SD and Delgado MK. Covid-19 related hospital admissions in the United States: needs and outcomes. BMJ 2020; 369: m2082. 2020/05/29. DOI: 10.1136/bmj.m2082.

8. Weissman GE, Crane-Droesch A, Chivers C, et al. Locally Informed Simulation to Predict Hospital Capacity Needs During the COVID-19 Pandemic. Ann Intern Med 2020; 173: 21–28. 2020/04/08. DOI: 10.7326/M20-1260.

9. World Health Organization. WHO R&D Blueprint-novel Coronavirus-COVID-19 Therapeutic Trial Synopsis, https://www.who.int/blueprint/priority-diseases/key-action/COVID-19_Treatment_Trial_Design_Master_Protocol_synopsis_Final_18022020.pdf (accessed 17 August 2020).

10. Singer M, Deutschman CS, Seymour CW, et al. The Third International Consensus Definitions for Sepsis and Septic Shock (Sepsis-3). JAMA 2016; 315: 801–810. 2016/02/24. DOI: 10.1001/jama.2016.0287.

11. Xu PP, Tian RH, Luo S, et al. Risk factors for adverse clinical outcomes with COVID-19 in China: a multicenter, retrospective, observational study. Theranostics 2020; 10: 6372–6383. 2020/06/03. DOI: 10.7150/thno.46833.

12. Goyal P, Choi JJ, Pinheiro LC, et al. Clinical Characteristics of Covid-19 in New York City. N Engl J Med 2020; 382: 2372–2374. 2020/04/18. DOI: 10.1056/NEJMc2010419.

13. Paliani U and Cardona A. COVID-19 and hydroxychloroquine: Is the wonder drug failing? Eur J Intern Med 2020 2020/06/20. DOI: 10.1016/j.ejim.2020.06.002.

14. Rivera-Izquierdo M, Del Carmen Valero-Ubierna M, JL Rd, et al. Sociodemographic, clinical and laboratory factors on admission associated with COVID-19 mortality in hospitalized patients: A retrospective observational study. PLoS One 2020; 15: e0235107. 2020/06/26. DOI: 10.1371/journal.pone.0235107.

15. Lechien JR, Chiesa-Estomba CM, Place S, et al. Clinical and epidemiological characteristics of 1420 European patients with mild-to-moderate coronavirus disease 2019. J Intern Med 2020 2020/05/01. DOI: 10.1111/joim.13089.

16. Rodriguez-Morales AJ, Cardona-Ospina JA, Gutierrez-Ocampo E, et al. Clinical, laboratory and imaging features of COVID-19: A systematic review and meta-analysis. Travel Med Infect Dis 2020; 34: 101623. 2020/03/18. DOI: 10.1016/j.tmaid.2020.101623.

17. Petrilli CM, Jones SA, Yang J, et al. Factors associated with hospital admission and critical illness among 5279 people with coronavirus disease 2019 in New York City: prospective cohort study. BMJ 2020; 369: m1966. 2020/05/24. DOI: 10.1136/bmj.m1966.

18. Bhargava A, Fukushima EA, Levine M, et al. Predictors for Severe COVID-19 Infection. Clin Infect Dis 2020 2020/05/31. DOI: 10.1093/cid/ciaa674.

19. Zheng Z, Peng F, Xu B, et al. Risk factors of critical & mortal COVID-19 cases: A systematic literature review and meta-analysis. J Infect 2020 2020/04/27. DOI: 10.1016/j.jinf.2020.04.021.

20. Turkiye Istatistik Kurumu (Turkish Institute of Statistics). Turkiye Saglik Arastirmasi (Health Report of Turkey), http://www.tuik.gov.tr/PreTablo.do?alt_id=1095 (accessed 17 August 2020).

21. Galloway JB, Norton S, Barker RD, et al. A clinical risk score to identify patients with COVID-19 at high risk of critical care admission or death: An observational cohort study. J Infect 2020 2020/06/02. DOI: 10.1016/j.jinf.2020.05.064.

22. Liang W, Liang H, Ou L, et al. Development and Validation of a Clinical Risk Score to Predict the Occurrence of Critical Illness in Hospitalized Patients With COVID-19. JAMA Intern Med 2020 2020/05/13. DOI: 10.1001/jamainternmed.2020.2033.

23. Cen Y, Chen X, Shen Y, et al. Risk factors for disease progression in patients with mild to moderate coronavirus disease 2019-a multi-centre observational study. Clin Microbiol Infect 2020 2020/06/12. DOI: 10.1016/j.cmi.2020.05.041.

24. Gong J, Ou J, Qiu X, et al. A Tool to Early Predict Severe Corona Virus Disease 2019 (COVID-19) : A Multicenter Study using the Risk Nomogram in Wuhan and Guangdong, China. Clin Infect Dis 2020 2020/04/17. DOI: 10.1093/cid/ciaa443.

25. Allenbach Y, Saadoun D, Maalouf G, et al. Multivariable prediction model of intensive care unit transfer and death: a French prospective cohort study of COVID-19 patients. medRxiv 2020: 2020.2005.2004.20090118. DOI: 10.1101/2020.05.04.20090118.

26. Barnes GD, Burnett A, Allen A, et al. Thromboembolism and anticoagulant therapy during the COVID-19 pandemic: interim clinical guidance from the anticoagulation forum. J Thromb Thrombolysis 2020; 50: 72–81. 2020/05/23. DOI: 10.1007/s11239-020-02138-z.

27. Korber B, Fischer W, Gnanakaran S, et al. Spike mutation pipeline reveals the emergence of a more transmissible form of SARS-CoV-2. bioRxiv 2020: 2020.2004.2029.069054. DOI: 10.1101/2020.04.29.069054.

28. Yao H, Lu X, Chen Q, et al. Patient-derived mutations impact pathogenicity of SARS-CoV-2. medRxiv 2020: 2020.2004.2014.20060160. DOI: 10.1101/2020.04.14.20060160.

